# Functional data for *LDLR* variant classification: comparative insights from high-content microscopy and flow cytometry assays

**DOI:** 10.64898/2026.05.09.26352817

**Authors:** Mohammad Majharul Islam, Ana Catarina Alves, Rafael Graça, Joana Rita Chora, Mafalda Bourbon, Simon G. Pfisterer

## Abstract

**Background and aims:** Current FH VCEP specifications of ACMG/AMP guidelines for familial hypercholesterolemia (FH) variant interpretation assign a higher evidence weight to functional data obtained with flow cytometry than microscopy assays, due to lack of existing evidence. This restricts the use of microscopy-derived functional data for variant classification. We aimed to systematically compare functional data of *LDLR* variants obtained by high-content microscopy and flow cytometry to determine their concordance and assess whether microscopy-based assays could support a higher evidence level.

**Methods:** Fifty *LDLR* variants with available flow cytometry and high-content microscopy data were compared for LDL uptake activity, including 21 newly characterized variants by microscopy in this study. Variants were grouped by FH VCEP functional thresholds (<70% activity, abnormal function; >90% activity, normal function) and results were integrated with UK Biobank data to assess associations with lipid traits.

**Results:** First, we validated our scalable microscopy assay with FH VCEP-classified control variants. Then we compared functional activity measured by microscopy and flow cytometry assays for 50 variants, which showed significant correlation (r = 0.66, p<0.0001) and a close average agreement (Bland–Altman bias = –0.05). Applying FH VCEP functional classification thresholds yielded broadly consistent classification in both methods, with minor shifts among categories. Integration with UK Biobank data showed that carriers of variants with reduced LDLR activity (<70% and <50%) had higher LDL-C, total cholesterol and ApoB levels compared to those with normal activity (>90%) for both microscopy and flow cytometry assays, with more pronounced differences observed at the <50% LDLR activity threshold.

**Conclusion:** High-content microscopy provides reliable and scalable measurements of LDLR function, showing high concordance with flow cytometry and consistent associations with lipid phenotypes. These findings support reconsideration of the evidence weight assigned to validated microscopy assays within FH VCEP variant classification frameworks, namely to Strong (Level 1).

## Introduction

Familial hypercholesterolemia (FH), an autosomal semi-dominant disorder characterized by markedly elevated LDL cholesterol (LDL-C) and an increased risk of premature atherosclerotic cardiovascular disease (ASCVD), represents one of the most common genetic disorders [1,2]. Whilst FH is underdiagnosed and undertreated in most countries [3], genetic testing holds great promise for early detection and effective management of the condition [4]. Low-density lipoprotein receptor (*LDLR*) gene variants are the primary genetic cause of FH [2,5] and advances in genetic testing have unraveled several thousand *LDLR* gene variants [6] (https://www.ncbi.nlm.nih.gov/clinvar/?gene=LDLR&term=%22LDLR%22%5BGENE%5D).

Accurate interpretation of the identified *LDLR* variants is essential for integration into clinical practice. However, a large fraction of variants lacking functional information hampers the classification of variants into benign or pathogenic [7,8]. Variant classifications by ClinGen Variant Curation Expert Panels (VCEP), such as the FH VCEP, are recognized by the Food and Drug Administration (FDA) as a trusted source for the interpretation of human genetic variants. This recognition allows ClinGen VCEP classifications to be used in clinical tests and in vitro diagnostics without requiring individual FDA review for each variant. As a result, FH VCEP-based classifications can directly influence clinical decision-making, including eligibility assessments and reimbursement decisions for lipid-lowering therapies [9]. However, the application of the ClinVar database is limited, as a substantial proportion of *LDLR* variants have not yet been classified by the FH VCEP, and the majority lack functional data, leading to many variants being classified as variants of uncertain significance (VUS). Assessing the functional consequences of *LDLR* variants on its expression, trafficking, and LDL uptake can provide mechanistic evidence for the disease phenotype and resolve uncertainty in variant classification. Accordingly, the American College of Medical Genetics and Genomics (ACMG) and the Association for Molecular Pathology (AMP) classification guidelines incorporate functional assay data as key evidence for variant interpretation, defined under the criteria PS3 and BS3 [8]. In the ClinGen – FH VCEP *LDLR* specifications these criteria are applied as following: PS3 (strong evidence for pathogenicity (<70% of wild-type activity) and BS3 (strong evidence for benign effect (>90% of wild-type activity)) [10,11].

Several methodological approaches have been implemented to quantify the impact of *LDLR* variants on receptor function. Among these, flow cytometry–based assays measuring fluorescently labeled LDL uptake in heterologous cell systems (e.g., *LDLR*-deficient Chinese Hamster Ovary (CHO) cells) have been widely adopted, generating functional evidence for a substantial number of *LDLR* variants [12–15]. In recent years, several novel microscopy-based methods have been published for the functional profiling of *LDLR* variants [16–19]. However, the ClinGen FH VCEP currently recognizes only flow cytometry–based assays as Level 1 (PS3/BS3 strong) functional evidence for *LDLR* variant classification, considering them as the reference methodology. In contrast, microscopy-based assays are classified as Level 3 (supporting) evidence, as there was insufficient evidence at the time to support their elevation to a higher evidence level [11]. This reflects the requirement for robust validation before such methods can be assigned higher evidentiary weight in variant classification.

Microscopy-based assays have the potential to directly examine how *LDLR* variants affect key steps in the receptor’s life cycle. We recently published a scalable, high-content fluorescence microscopy platform for the functional profiling of *LDLR* variants [17]. The system utilizes a CRISPR-engineered *LDLR*-deficient liver cell model (HepG2) and automated confocal microscopy to quantitatively assess receptor expression and DiI-LDL internalization at single-cell resolution. Importantly, by linking this functional data to clinical phenotypes from the UK Biobank, we demonstrated that the residual activity of *LDLR* variants stratifies plasma lipid concentrations and cardiovascular risk in an activity-dependent gradient [17].

To provide further evidence that microscopy assays can be used equivalently for the functional characterization of *LDLR* variants, we performed a systematic comparative analysis between flow cytometry- and microscopy-based functional profiling across 50 *LDLR* variants, including 21 newly analyzed variants with the microscopy assay. This study benchmarks our analysis platform and provides support for raising the evidence level for automated microscopy assays in future *LDLR* variant classification guidelines.

## Materials and Methods

### Reagents

Hoechst 33342 (Thermo Fisher, Cat: H1399), CellMask Deep Red (Thermo Fisher, Lot: 216004), Lipofectamine LTX & PLUS reagents (Cat: 15338030), Dil Stain (1,1’-Dioctadecyl-3,3,3’,3’-Tetramethylindocarbocyanine Perchlorate (’DiI’; DiIC18(3))) (Cat: D3911) and fetal bovine serum (FBS) (Cat: 10270106) were purchased from Thermo Fisher. Q5 Hot Start High-Fidelity DNA polymerase (Cat: M04932), Phusion High Fidelity Polymerase (Cat: F530S), Taq DNA ligase (Cat: M0208S), T5 Exonuclease (Cat: M0363S), Bgl II (Cat: ER0081), EcoRI-HF (Cat: R3101S) were purchased from New England Biolabs. L-glutamate (Cat: BE17-605E) and Penicillin streptomycin (Cat: DE17-602E) from Lonza. Trypsin/EDTA solution (Cat: R-001-100) from gibco, Poly-L-lysine (Cat: 11426642) from MP Biochemicals, RPMI-1640 (Cat: ECB9006L) from EuoClone, SeraSil-Mag 400 silica coated superparamagnetic beads (Cat: 29357371) from Cytiva, and dNTPs (Cat: U1240) were purchased from Promega. Lipoprotein-deficient serum (LPDS), DiI-labeled low-density lipoprotein (DiI-LDL) were produced as previously described (Reynolds and St. Clair 1985; Stephan and Yurachek 1993).

### Cell culture

HepG2 cells were obtained from the European Collection of Authenticated Cell Cultures (ECACC). The *LDLR* knockout HepG2 cell line was generated using CRISPR-Cas9 technology as previously described [17]. HepG2 and *LDLR* knockout HepG2 cells were maintained in a complete cell culture medium consisting of RPMI-1640 medium supplemented with 10% fetal bovine serum (FBS), 1% L-glutamine, and 1% penicillin–streptomycin at 37° C and 5% CO2 in a humidified incubator. *LDLR* knockout cells stably expressing *LDLR* variants were maintained in cell culture medium. Cells expressing *LDLR* variants in an *LDLR* knockout background were maintained in complete culture medium supplemented with 2 µg/mL puromycin. Cells were subcultured upon reaching 80– 90% confluence using 0.025% Trypsin/EDTA solution.

### Variant Selection

A panel of *LDLR* variants functionally characterized using flow cytometry was curated from previously published peer-reviewed articles [12–15,20–35]. Variants were included based on the availability of quantitative measurements of LDL uptake and LDLR expression. Intersection of this panel with the variants analyzed using our microscopy-based platform identified 29 shared variants, which were used for comparative analysis as well as additional 21 variants that were newly analyzed for the present study. LDL uptake measurements for the curated variants are summarized in Supplementary Table 1.

To validate the microscopy-based LDL uptake assay, we first analyzed a control set of 25 *LDLR* variants with established clinical classifications. These included 20 pathogenic and five benign/likely benign variants curated and classified by the FH VCEP, a ClinGen expert panel recognized by FDA for variant interpretation.

### Variant construction and functional assay

The detailed methodology for *LDLR* variant construction and functional analysis using a microscopy-based platform has been described previously [17]. Briefly, LDLR variant expression constructs were generated using a semi-automated robotic platform. For each variant, the *LDLR* gene was amplified in two fragments, with the desired mutation introduced via oligonucleotides. These fragments were then assembled into the linearized GFP-tagged expression vector (pSH-EFIRES-P) using Gibson assembly [36], generating an LDLR variant expression construct in which GFP is fused to the C-terminus of the LDLR protein. All variant constructs were verified by next-generation sequencing. Stable LDLR variant–expressing HepG2 cell lines were generated by integrating the variant expression cassette into the AAVS1 safe-harbor locus using CRISPR/Cas9 technology.

Functional characterization of each *LDLR* variant was performed using a fluorescent low-density lipoprotein (DiI-LDL) uptake assay. Variant-expressing cell lines were seeded in 384-well plates and cultured under lipid-rich (RPMI medium supplemented with 10% FBS) or lipid-poor conditions (serum-free medium supplemented with 5% LPDS). For LDL uptake, cells were incubated with 10 µg/mL DiI-LDL for 30 minutes at 37 °C. Following incubation, cells were fixed, stained with Hoechst 33342 and CellMask Deep Red, and imaged using a PerkinElmer Opera Phenix high-content spinning-disk confocal microscope with a 20x objective.

Each variant was analyzed in triplicate wells across two independent experiments, with approximately 2,000 cells quantified per variant cell line. Image analysis was performed using CellProfiler to quantify functional readouts, including the mean cellular DiI-LDL intensity, the number of DiI-LDL-positive organelles per cell, and mean cellular variant expression. These readouts were normalized to wild-type LDLR-GFP controls to determine variant-specific receptor activity. Because variants in the ligand-binding domain exhibited increased receptor expression, an additional correction step was applied to adjust their activity scores for overexpression-related biases.

### UK Biobank data

We leveraged whole-exome sequence (WES) data from the UK Biobank to extract clinical data for carriers of specific *LDLR* variants. Among the 50 variants included in our comparative analysis, 33 were identified in 8773 UK Biobank participants. Lipid profile data for variant carriers derived from the first UK Biobank assessment was used for analysis. Genetic and phenotypic data were obtained through the UK Biobank Research Analysis Platform (RAP).

### Statistical analysis

Statistical analyses were conducted in Python using pandas, numpy, scipy, matplotlib, and seaborn. Correlation between the microscopy- and flow cytometry–based functional readouts was quantified using Spearman’s correlation, and agreement between assay readouts was evaluated through Bland– Altman analysis. Pairwise comparisons between variant activity groups were performed using the 2-sided Mann-Whitney U test. Bonferroni adjustments were applied for multiple testing.

## Results

### Validation of the microscopy-based assay and comparison with flow cytometry

To validate the microscopy-based LDL uptake assay, we first evaluated a control set of 25 *LDLR* variants previously classified by the FH VCEP as benign or pathogenic. All five benign variants demonstrated >90% LDL uptake activity in the microscopy assay, including two variants in which functional data was considered for variant classification (Fig. 1A, green checkered bars) and three variants without previous functional evidence (Fig. 1A, green solid bars). Of the 20 variants classified as pathogenic by FH VCEP, 12 variants included functional data in their evaluation. For these 12 pathogenic variants, the microscopy assay provided residual LDLR activities within the 0-70% activity range, concordant with the FH VCEP functional thresholds for pathogenic classification (Fig. 1A, red checkered bars). The remaining eight variants were classified as pathogenic without considering functional data (Fig. 1A, red solid bars). Of these eight variants four remained well below the 70% activity threshold, three variants were within the 70-90% activity range and one variant (c.1897C>T/p.R633C) displayed wild-type-like activity (>90% activity).

**Figure 1:**
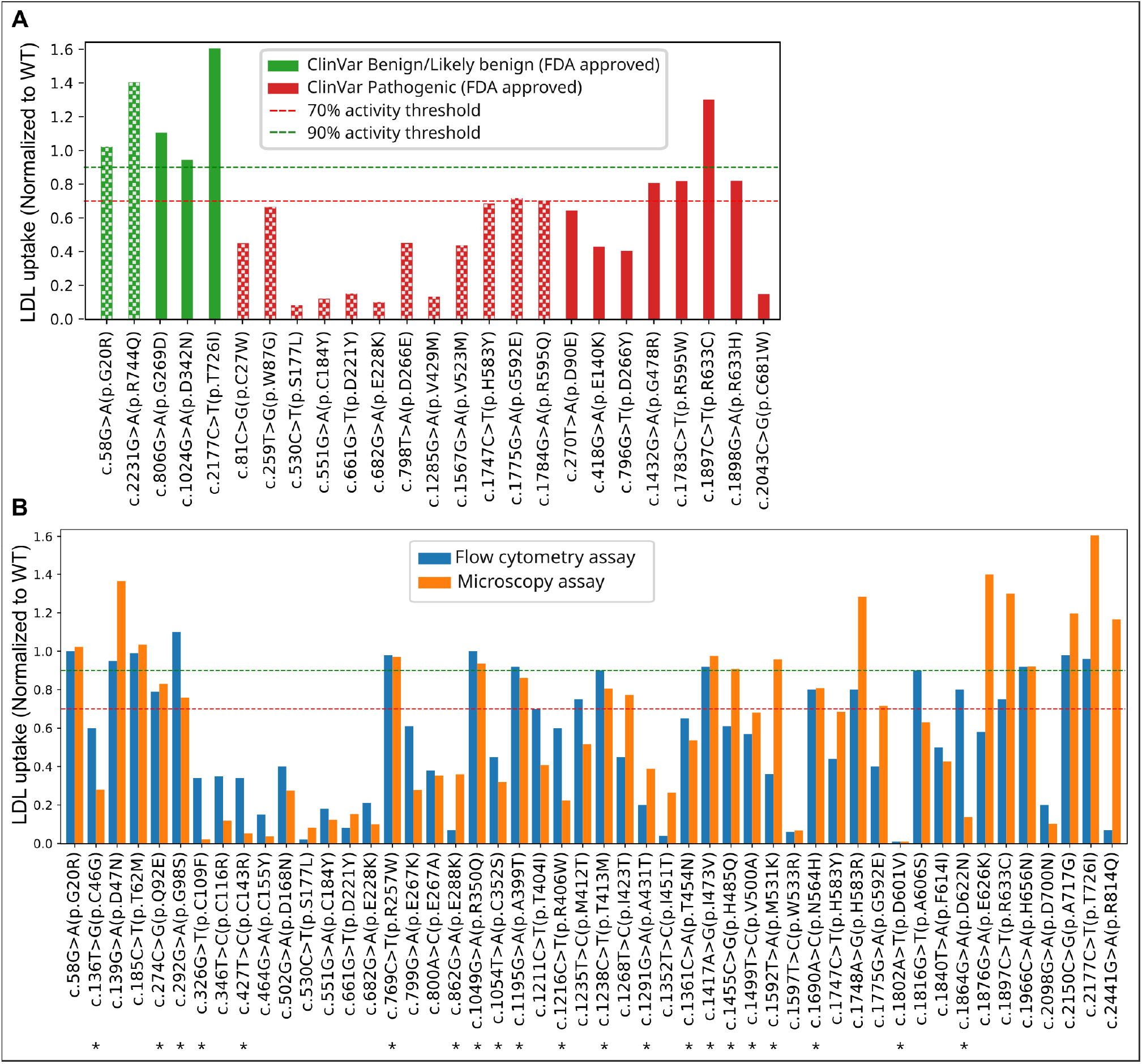
Functional characterization of *LDLR* variants by microscopy- and flow cytometry-based assays. (A) Validation of our semi-automated microscopy-based functional assay using a control set of variants, comprising five LDLR variants classified as benign/likely benign and 20 variants classified as pathogenic by the ClinVar expert panel. Checkered patterns indicate variants for which functional data contributed to the classification process (B) Comparison of LDL uptake activities for all analyzed LDLR variants as measured by microscopy (orange bars) and flow cytometry (blue bars). Microscopy-derived functional scores represent the mean wild-type normalized values from two independent experiments performed in triplicate. Flow cytometry–derived LDL uptake values for each variant were obtained from previously published data. Dashed green and red lines indicate 90% and 70% activity thresholds, respectively. Asterisks (*) indicate variants newly analyzed in this study using the microscopy-based assay.

Next, we selected 50 *LDLR* variants previously characterized with flow-cytometry and compared their LDL uptake activity with microscopy data for the same variants (Fig. 1B). Of these, functional microscopy data for 29 variants had been reported previously [17], while the remaining 21 variants were newly analyzed using the microscopy platform in this study (indicated by * in Fig. 1B). Among these newly characterized variants, 11 showed <70% activity and 10 variants showed >90% activity.

To further examine the alignment between microscopy and flow cytometry-based assays, we performed correlation and agreement analyses of LDL uptake values. Correlation analysis demonstrated a significant positive relationship (r = 0.66, p = 1.28 × 10^−7^) between microscopy- and flow cytometry-derived measurements, indicating that both methods capture similar trends across variants (Fig. 2A). Agreement analysis using a Bland–Altman plot demonstrated minimal systematic bias between the two methods (bias = –0.05), with most variants falling within the 95% limits of agreement (Fig. 2B). While variant-level discrepancies were observed, the overall distribution supported good concordance between microscopy- and flow cytometry-based LDL uptake measurements.

**Figure 2:**
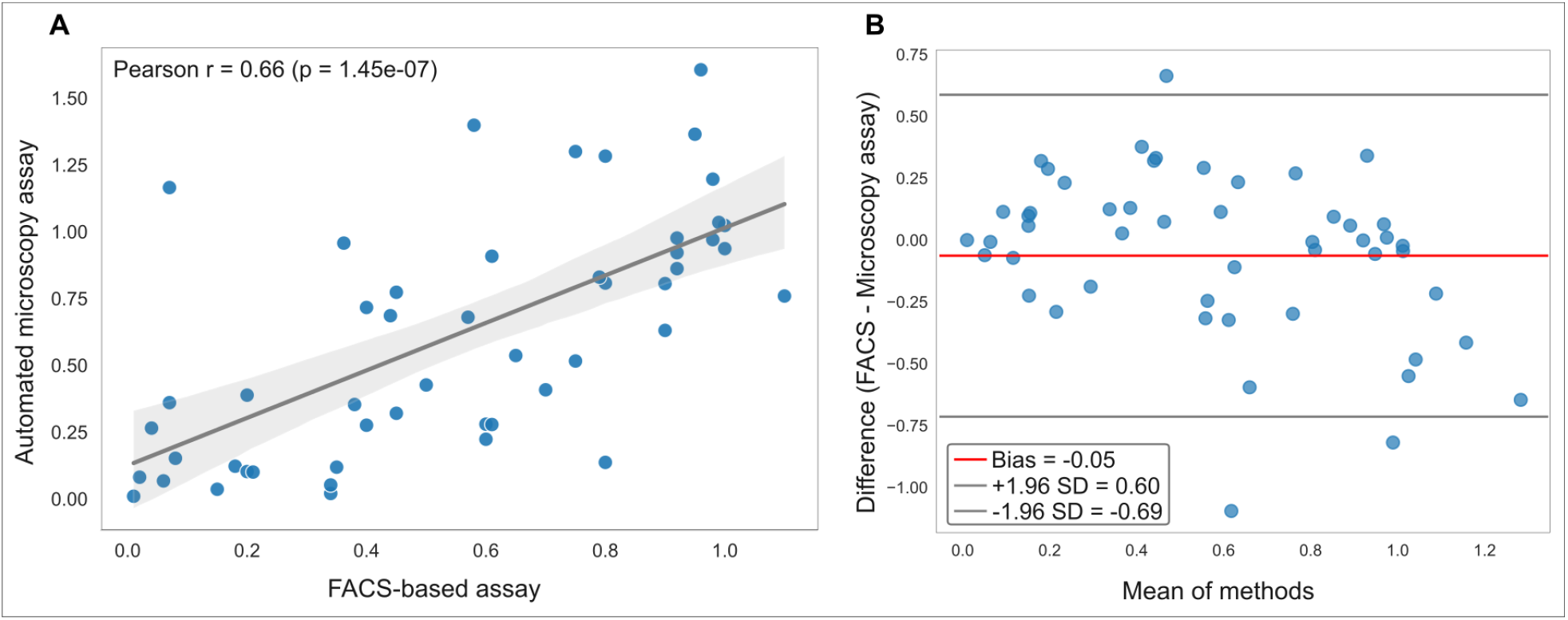
Correlation and agreement between microscopy-based and flow cytometry-based *LDLR* activity measurements. (A) Correlation analysis showing a significant positive relationship among LDL uptake scores measured by two methods (r = 0.66, p = 1.45 × 10^−7^). The shaded area indicates the 95% confidence interval of the regression line. (B) Bland-Altman plot assessing agreement between the two methods. The red line indicates the mean difference (bias) of -0.05, indicating a minimal average difference between microscopy and flow cytometry measurements. The grey lines represent the 95% limits of agreement, showing the range within which most differences between the two methods fall. Each blue circle represents a single LDLR variant.

### Categorization of *LDLR* variants using functional data from microscopy and flow cytometry assays

Using LDL uptake values measured by both assays, we next categorized variants into functional groups according to FH VCEP guidelines: defective (<70% LDL uptake), grey zone (70–90% LDL uptake), and wild-type-like (>90% LDL uptake) (Fig. 3). With microscopy-derived data, 28 of the 50 variants (56%) were categorized as defective, 15/50 variants (30%) as wild-type-like, and 7/50 variants (14%) as grey zone (Fig. 3A). In comparison, slightly more variants were grouped as defective with the flow cytometry assay (30/50, 60%) and slightly fewer as wild-type-like (11/50, 22%) (Fig. 3B).

**Figure 3:**
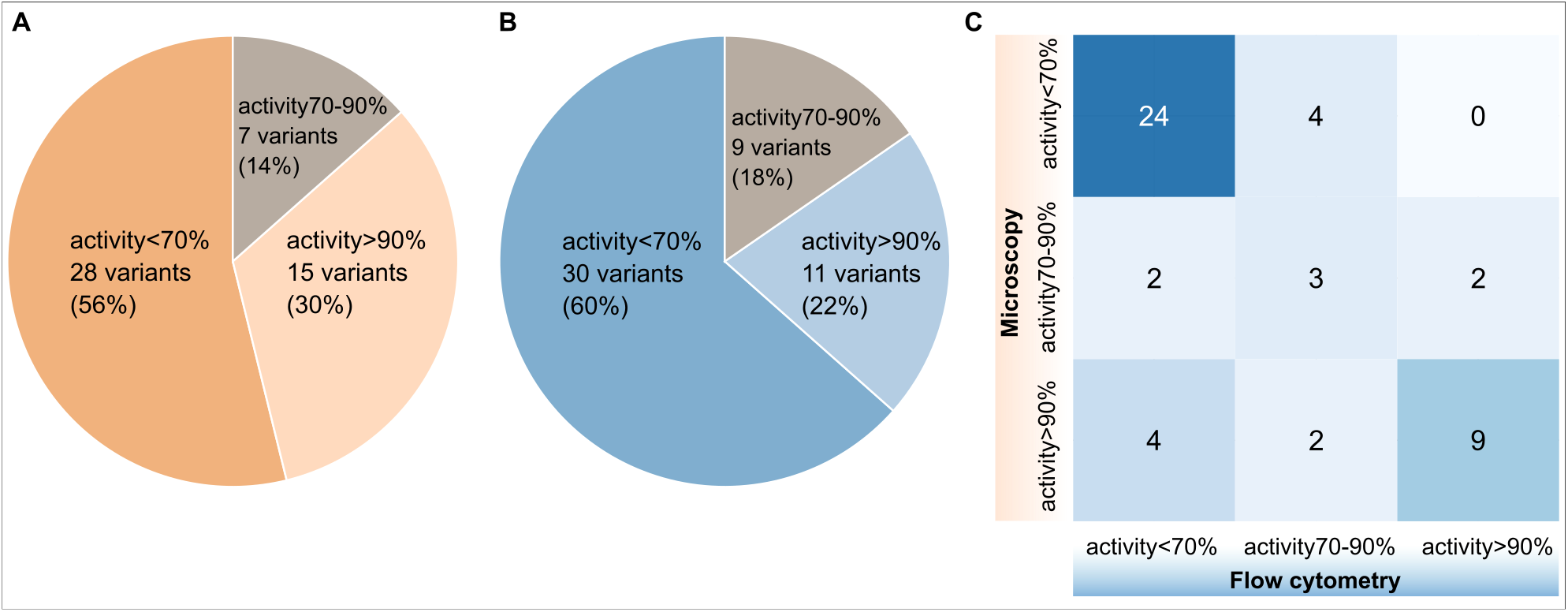
Categorization of *LDLR* variants by functional activity measured by microscopy-based and flow cytometry-based assays. Variants were classified into three activity groups according to FH Variant Curation Expert Panel guidelines: activity < 70%, activity 70–90%, and activity > 90%, based on LDL uptake activity. (A) Distribution of variants by activity category using microscopy-based LDL uptake data. (B) Distribution of variants by activity category using flow cytometry-based LDL uptake assay results. (C) Contingency table showing the overlap in variant classification between the two methods, highlighting concordance and discordance across activity categories.

Direct comparison of categorical assignments by the two methods showed modest overall concordance (35/50, 70.0%) but also revealed shifts between groups (Fig. 3C). Of the 35 variants which agree in functional categorization across both methods, 24 were classified as defective (<70%), three were categorized into the grey zone (70-90%) and nine into the wild-type-like (>90%) group for both methods. Importantly, 85.7% (24/28) of variants grouped as defective by microscopy assays were also classified as defective using LDL uptake data derived from flow cytometry assays. The remaining 14.3% (4/28) of microscopy-assigned defective variants were categorized as grey zone (70-90) by flow-cytometry data, with none shifting to the wild-type-like category. 60% (9/15) of variants assigned as wild-type-like based on microscopy assays were also wild-type-like with flow cytometry-derived LDL uptake data. Out of the six microscopy-assigned wild-type-like variants that disagreed with flow cytometry categorization, four (26.7%) were categorized as defective and two (13.3%) as grey zone when categorized based on flow cytometry-derived functional data. Considering the flow cytometry categorization, variants largely retained the same categories assigned by microscopy categorization: 81.8% (9/11) remaining wild-type-like and 80% (24/30) remaining defective across both methods. For both assays, most shifts were observed in the grey zone category, reflecting its activity range (70-90%) near the threshold boundaries (Fig. 3C). Of the seven variants categorized as grey zone by microscopy assay data, two shifted to the defective group and two to the wild-type-like groups obtained by flow cytometry data. Regarding the nine variants categorized as grey zone by flow cytometry data, four moved to the defective category and two shifted to the wild-type-like group when categorization was performed using microscopy-derived data (Fig. 3C).

### Relationship of functional activity groups with blood lipid levels in UK Biobank

Previously we demonstrated the effects of variant LDL uptake groups on plasma lipid levels by integrating the functional data with WES data for 500 000 subjects in UK Biobank. In this study, we employed this strategy to better evaluate the comparability of flow-cytometry and microscopy LDL uptake data regarding its clinical relevance at the population level. Therefore, utilizing the WES data, UK Biobank participants carrying the *LDLR* variants analyzed in this study were extracted along with their plasma lipid measurements. Participants carrying variants within the <70% and >90% activity groups were then selected for association analysis. The same association studies were performed with a more stringent <50% activity subgroup to further assess the clinical impact of more severe functional impairment. Only individuals who did not receive lipid-lowering medications were included in the analysis. The number of variants and carriers in each functional activity group, for both assays, is summarized in Fig. 4A.

**Figure 4:**
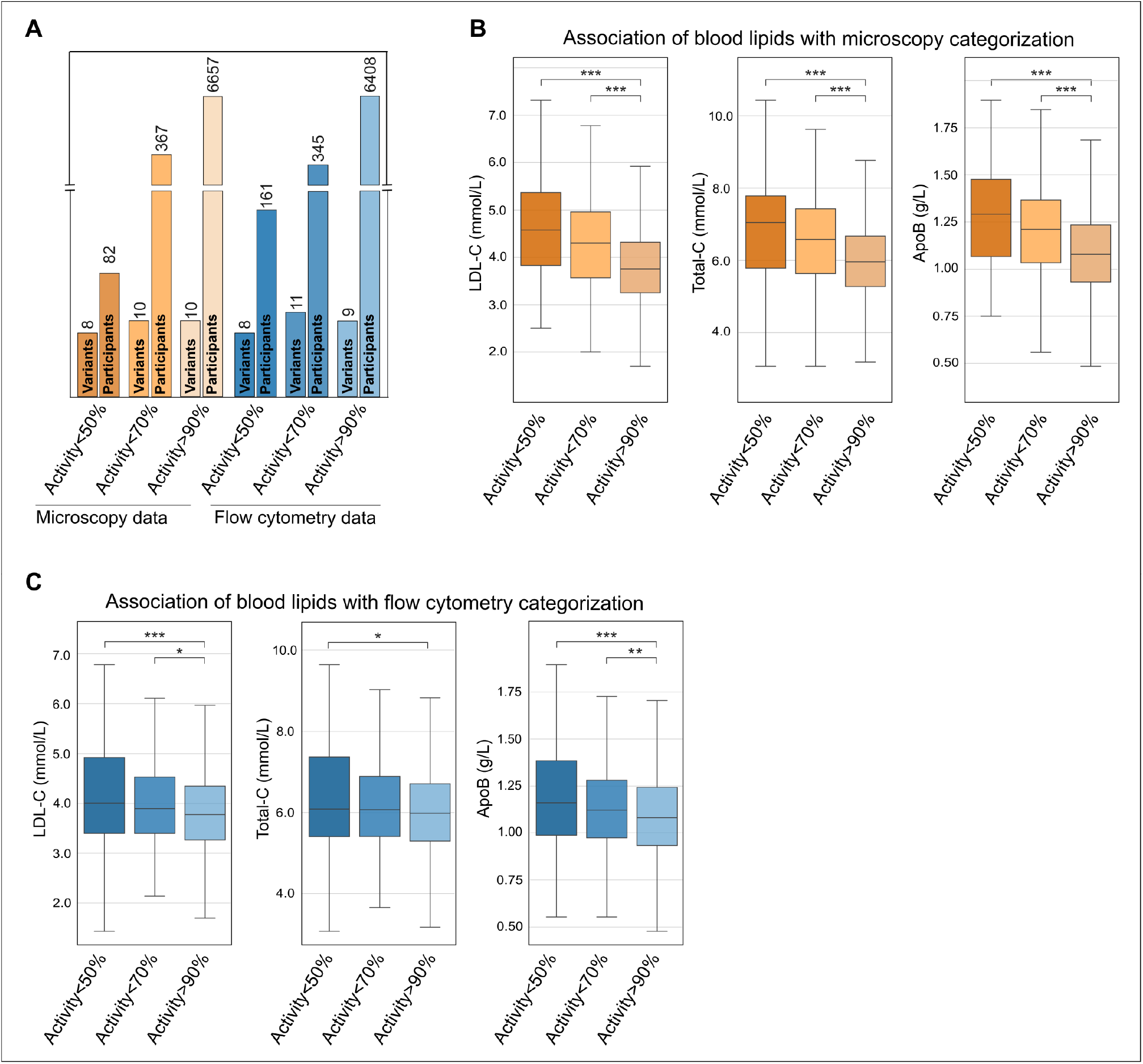
Association of *LDLR* variant activity categories with blood lipid concentrations in UK Biobank participants. (A) Bar chart displays the number of LDLR variants and corresponding UK Biobank carrier counts for each functional activity category. LDL cholesterol (LDL-C), Total cholesterol (Total-C), and Apolipoprotein B (ApoB) levels in nontreated individuals are consistently higher in carriers of low-activity variants (<50% and <70%) compared with high-activity variants (>90%), for both (B) Microscopy and (C) Flow cytometry methods. Data were analyzed using the Mann-Whitney U test. P values were corrected for multiple pair tests. Asterisks denote significance levels after Bonferroni correction: * P < 0.05, ** P < 0.01, and *** P < 0.001.

Carriers of variants in both microscopy and flow cytometry-based <70% groups displayed higher mean LDL-C (MIC: 4.3 vs 3.81 mmol/L, FLOW: 3.99 vs 3.81 mmol/L), total cholesterol (MIC: 6.58 vs 6.01 mmol/L, FLOW: 6.19 vs 6.01 mmol/L) and apolipoprotein B (MIC: 1.22 vs 1.09 g/L, FLOW: 1.14 vs 1.09 g/L) compared with the >90% activity group (Fig. 4B-C). The effects were more pronounced when comparing the <50% group with the >90% groups for both flow cytometry and microscopy assays for LDL-C (MIC: 4.57 vs 3.81 mmol/L, FLOW: 4.18 vs 3.81 mmol/L), total cholesterol (MIC: 6.88 vs 6.01 mmol/L, FLOW: 6.38 vs 6.01 mmol/L) and ApoB (MIC: 1.3 vs 1.09 g/L, FLOW: 1.19 vs 1.09 g/L) (Fig. 4B-C). No significant differences were observed in serum triglyceride between groups defined by either assay.

## Discussion

Functional profiling of *LDLR* variants is critical for FH diagnosis as it improves our understanding of their functional impact on cellular disease mechanisms. Integration of functional evidence expands the data available for variant classification by the FH VCEP, thereby increasing the number of variants that can be confidently classified as disease-causing or not. The main goal of this study was to enable improved utilization of scalable microscopy assays for variant classification by the FH VCEP. This will enable more individuals living with FH to receive a definitive diagnosis, facilitating clinical decision-making, and improved management of the disease.

Building on the original radiolabeled assays, functional analysis of *LDLR* variants has increasingly relied on flow cytometry and microscopy platforms, which quantify fluorescent LDL uptake in parallel with receptor expression and intracellular trafficking, thereby providing mechanistic insights into FH pathogenicity. However, a direct comparative evaluation between microscopy and flow cytometry approaches has been lacking. This study aimed to evaluate the performance of a semi-automated microscopy assay, previously applied by our group to characterize 315 *LDLR* variants [17], with the goal to scale to most *LDLR* missense variants in the near future. Demonstrating comparability between microscopy and flow cytometry assays would provide key validation supporting the use of microscopy-derived data in FH VCEP variant classification frameworks.

Direct comparison of LDL uptake values revealed a significant correlation and overall agreement between microscopy- and flow cytometry-based assays, supporting the reliability of both approaches for quantifying receptor activity. Accordingly, functional categories derived from microscopy overlapped largely with those from flow cytometry when FH VCEP cutoffs were applied. It is noteworthy to mention that the flow-cytometry data have been collected from studies performed in different laboratories. Moreover, the methodologies are different, in flow cytometry surface-bound LDL is quenched with trypan blue to detect internalized LDL, whilst in the microscopy assay overall cellular intensity and accumulation of fluorescent LDL particles in endosomes is quantified. Furthermore, different cell systems have been used. Whilst the microscopy assays are carried out with an *LDLR* CRISPR knockout liver cell line in which the LDLR fused to green fluorescent protein is reintroduced, the flow cytometry assays were mostly performed with *LDLR* deficient CHO cells. Overall, LDL uptake data from microscopy assays largely reflect the observations obtained with the flow cytometry assay, and discrepancies for a small subset of variants are likely attributable to methodological and cell system–specific factors. A striking discrepancy was observed for variant c.2441G>A (p.R814Q), with the microscopy-based analysis reporting wild-type-like activity, whereas the flow cytometry assay measured severely impaired activity [34]. Since both assays utilized a HepG2 cell model, cell-type-specific effects are unlikely to account for the contrasting results. However, Mori et al. first introduced a double strand break with CRISPR/Cas9 and then utilized homology directed repair to introduce the gene variant. Usually only a subset of cells undergoes homology directed repair, requiring single cell cloning and validation of the correct insertion of the gene variant. However, only cell pools, not cells enriched for the variant p.R814Q, were used for downstream analysis. Moreover, the integration of p.R814Q was not validated by sequencing. This may indicate that the results are mostly derived from HepG2 *LDLR* knockout cells rather than a cell system representing the p.R814Q variant. This could explain the observed discrepancies in residual LDL uptake activity with our microscopy assay. In line with our microscopy-based characterization, another flow cytometry-based functional screening study, in which a genetic variant resulting in the same amino acid change (p.R814Q) was expressed from a cDNA construct in HeLa cells, reported wild-type–like activity (LDL uptake 104%, surface expression 102% of WT [37].

When comparing our microscopy results with ClinGen FH VCEP classification, one *LDLR* variant (c.1897C>T (p.R633C)) displayed discordant results with our microscopy functional characterization. In our study, the variant exhibited wild-type-like receptor function. However, the ClinGen FH VCEP classifies this variant as pathogenic based on multiple entries with genetic and clinical evidence. The variant has been identified in at least 10 unrelated index cases across global populations who meet stringent FH clinical criteria and shows segregation with the phenotype in two distinct families. Additional criteria for pathogenic classification comes from its low allele frequency (MAF ∼0.003%) and presence of another pathogenic variants affecting the same codon (p.R633H). Notably, previous studies have also reported mixed functional data for this variant, with one study showing reduced activity (∼75%) [14], whereas another study reported wild-type-like activity (107%) [16]. Given that a large set of clinically defined FH patients does not contain disease causing FH gene variants, it remains open whether clinical variant classification or functional studies are representing the real variant effect. All functional studies for this variant were performed with ectopically expressed *LDLR* variant cDNA. Therefore, it could be possible that localization in the native genomic environment affects the expression of variant p.R633C and LDLR activity. It may be relevant to investigate LDLR expression or LDL uptake in primary patient cells to better understand the functional consequences of this variant.

The relevance of microscopy- and flow cytometry-derived functional data was highlighted with our UK Biobank integration, which demonstrated that for both assay procedures variants with reduced activity resulted in elevated LDL-C, ApoB, and total cholesterol levels as compared to variants with a residual activity above 90%. The differences were more pronounced when comparing the <50% group with the >90% group. This reflects previous observations that the effect of variants on LDL-C with a residual activity of 30-70% is lower as compared to *LDLR* variants with 0-30% residual activity [17]. Potentially, these results have interesting implications on the use of functional data in FH VCEP classification. It may suggest that the threshold for residual LDLR activity could be re-evaluated or a graded evidence level applied, to better reflect the functional activity of a variant. However, this question is complex as mildly defective variants can be relatively common, increasing the possibility that a person carries two of such variants. More functional data will enable smaller LDLR activity groups, providing more fine grained effects on blood lipids and CVD risk. In the future, such data will be valuable for the interpretation and classification of *LDLR* variants.

Interestingly, the defective *LDLR* variant group defined by microscopy data exhibited greater differences in LDL-C than the corresponding group defined by flow cytometry. This discrepancy may reflect the use of human liver-derived HepG2 cells in microscopy assays. Approximately 90% of variants in flow cytometry-based assays were evaluated in CHO cells, a non-hepatic, non-human system. The absence of the native hepatic interactome in CHO cells may alter protein-protein interactions and trafficking dynamics, particularly for variants involving single amino acid substitutions. As a result, CHO cells may overestimate functional impairment of *LDLR* variants, categorizing more variants as defective compared to HepG2 cell-based assays, thereby compromising the discrimination in plasma lipid levels between defective and non-defective variant carriers.

Whilst flow cytometry and microscopy assays provide comparable functional data for *LDLR* variants, the existing high-content based analysis pipeline provides several additional advantages: 1) Stable genomic integration of the LDLR variant expression constructs ensures low-level and homogeneous protein expression, enabling precise quantification of receptor expression; 2) Utilization of stable LDLR variant cell lines enables quantification of a large pool of cells, overcoming intercellular variability and making it easier to provide a graded LDLR activity; 3) The modularity of the high-content imaging approach enables extension to other readouts and gene variants.

This study is limited by the number of *LDLR* variants with available paired functional data and by the use of observational lipid phenotypes, although the consistency across assays and clinical datasets supports the robustness of the findings.

## Conclusion

We present a systematic comparison of a novel high-content microscopy assay with established flow cytometry method for functional characterization of *LDLR* variants. The functional data of *LDLR* variants derived from the microscopy-based assay showed high concordance with flow cytometry results and demonstrated a clear alignment with clinical phenotypes in individuals carrying these variants. In addition, the semi-automated microscopy platform offers the advantage of higher throughput, enabling the efficient analysis of large numbers of *LDLR* variants and variants in other FH-associated genes. Overall, our results strongly support a reassessment of the FH VCEP guidelines to elevate this HepG2 cell-based microscopy assay from Level 3 (Supporting) to Level 1 (Strong) evidence. This upgrade will significantly strengthen *LDLR* variant classification and improve FH management.

## Data Availability

All data produced in the present study are available upon reasonable request to the authors

## Author contributions

Mohammad Majharul Islam performed and analyzed the microscopy-based functional characterization of *LDLR* variants, integrated UK Biobank data with the functional results, conducted the statistical analyses, and wrote the manuscript. Ana Catarina Alves and Rafael Graça curated the flow cytometry-based functional data for the *LDLR* variants and reviewed the manuscript. Joana Rita Chora reviewed and validated the classification of *LDLR* variants included in this study and reviewed the manuscript. Mafalda Bourbon contributed to the conceptualization of the manuscript and provided important input through review and feedback. Simon G. Pfisterer supervised the functional study of *LDLR* variants and data analysis, as well as contributed to experimental design, conceptualization and editing of the manuscript.

## Acknowledgement

This research has been conducted using the UK Biobank resource under application number 87446.

