## Supplementary Table for "Functional data for *LDLR* variant classification: comparative insights from high-content microscopy and flow cytometry assays"

**Supplementary Table 1:** LDL uptake measurements by flow cytometry and microscopy assays

| *LDLR* variants | LDL uptake_flow cytometry | LDL uptake_microscopy | References(flow cytometry; microscopy) |
|---|---|---|---|
| c.136T>G (p.C46G) | 0.6 | 0.28 | [1]; This study |
| c.274C>G (p.Q92E) | 0.79 | 0.83 | [2]; This study |
| c.292G>A (p.G98S) | 1.1 | 0.76 | [3]; This study |
| c.326G>T (p.C109F) | 0.34 | 0.02 | [4]; This study |
| c.427T>C (p.C143R) | 0.34 | 0.05 | [4]; This study |
| c.769C>T (p.R257W) | 0.98 | 0.97 | [5]; This study |
| c.862G>A (p.E288K) | 0.07 | 0.36 | [6]; This study |
| c.1049G>A (p.R350Q) | 1 | 0.94 | [7]; This study |
| c.1054T>A (p.C352S) | 0.45 | 0.32 | [4]; This study |
| c.1195G>A (p.A399T) | 0.92 | 0.86 | [8]; This study |
| c.1216C>T (p.R406W) | 0.6 | 0.22 | [9]; This study |
| [2]c.1238C>T (p.T413M) | 0.9 | 0.81 | [8]; This study |
| c.1291G>A (p.A431T) | 0.2 | 0.39 | [10]; This study |
| c.1361C>A (p.T454N) | 0.65 | 0.54 | [11]; This study |
| c.1417A>G (p.I473V) | 0.92 | 0.98 | [2]; This study |
| c.1455C>G (p.H485Q) | 0.61 | 0.91 | [4]; This study |
| c.1499T>C (p.V500A) | 0.57 | 0.68 | [4]; This study |
| c.1592T>A (p.M531K) | 0.36 | 0.96 | [12]; This study |
| c.1690A>C (p.N564H) | 0.8 | 0.81 | [13]; This study |
| c.1802A>T (p.D601V) | 0.01 | 0.01 | [2]; This study |
| c.1864G>A (p.D622N) | 0.8 | 0.14 | [3]; This study |
| c.1235T>C (p.M412T) | 0.75 | 0.52 | [14]; [15] |
| c.2150C>G (p.A717G) | 0.98 | 1.2 | [10]; [15] |
| c.346T>C (p.C116R) | 0.35 | 0.12 | [5]; [15] |
| c.551G>A (p.C184Y) | 0.18 | 0.12 | [2]; [15] |
| c.1966C>A (p.H656N) | 0.92 | 0.92 | [2]; [15] |
| c.1211C>T (p.T404I) | 0.7 | 0.41 | [16]; [15] |
| c.1597T>C (p.W533R) | 0.06 | 0.07 | [12]; 15] |
| c.799G>A (p.E267K) | 0.61 | 0.28 | [4]; [15] |
| c.1816G>T (p.A606S) | 0.9 | 0.63 | [2][15] |
| c.464G>A (p.C155Y) | 0.15 | 0.04 | [11][15] |

| | | | |
|---|---|---|---|
| c.502G>A (p.D168N) | 0.4 | 0.28 | [5][15] |
| c.661G>T (p.D221Y) | 0.08 | 0.15 | [2][15] |
| c.139G>A (p.D47N) | 0.95 | 1.36 | [17]; [15] |
| c.2098G>A (p.D700N) | 0.2 | 0.1 | [18]; [15] |
| c.682G>A (p.E228K) | 0.21 | 0.1 | [10]; [15] |
| c.800A>C (p.E267A) | 0.38 | 0.35 | [7]; [15] |
| c.1876G>A (p.E626K) | 0.58 | 1.4 | [4]; [15] |
| c.1840T>A (p.F614I) | 0.5 | 0.43 | [4][15] |
| c.58G>A (p.G20R) | 1 | 1.02 | [2]; [15] |
| c.1775G>A (p.G592E) | 0.4 | 0.72 | [2]; [15] |
| c.1748A>G (p.H583R) | 0.8 | 1.28 | [19]; [15] |
| c.1747C>T (p.H583Y) | 0.44 | 0.69 | [19]; [15] |
| c.1268T>C (p.I423T) | 0.45 | 0.77 | [10]; [15] |
| c.1352T>C (p.I451T) | 0.04 | 0.27 | [4]; [15] |
| c.1897C>T (p.R633C) | 0.75 | 1.3 | [8]; [15] |
| c.2441G>A (p.R814Q) | 0.07 | 1.17 | [20]; [15] |
| c.530C>T (p.S177L) | 0.02 | 0.08 | [21]; [15] |
| c.185C>T (p.T62M) | 0.99 | 1.03 | [17]; [15] |
| c.2177C>T (p.T726I) | 0.96 | 1.61 | [2]; [15] |

Note: LDL uptake measurements for the variants are normalized to wild-type LDLR activity.
